# Staff to staff transmission as a driver of healthcare worker infections with COVID-19

**DOI:** 10.1101/2020.12.25.20248824

**Authors:** Claire L Gordon, Jason A Trubiano, Natasha E Holmes, Kyra YL Chua, Jeff Feldman, Greg Young, Norelle L Sherry, M Lindsay Grayson, Jason C Kwong

**Author notes:** **Corresponding author:** Dr Jason Kwong –.

## Abstract

**Objectives:** To investigate the COVID-19 infections among staff at our institution and determine the interventions required to prevent subsequent staff infections.

**Design:** Retrospective cohort study

**Participants and setting:** Staff working at a single tertiary referral hospital who returned a positive test result for SARS-CoV-2 between 25 January 2020 and 25 November 2020.

**Main outcome measures:** Source of COVID-19 infection.

**Results:** Of 45 staff who returned a positive test result for SARS-CoV-2, 19 were determined to be acquired at Austin Health. Fifteen (15/19; 79% [95% CI: 54–94%]) of these were identified through contact tracing and testing following exposures to other infected staff and were presumed to be staff-staff transmission, including 10 healthcare workers (HCWs) linked to a single ward that cared for COVID-19 patients. Investigation of the outbreak identified the staff tearoom as the likely location for transmission, with subsequent reduction in HCW infections and resolution of the outbreak following implementation of enhanced control measures in tearoom facilities. No HCW contacts (0/204; 0% [95% CI: 0–2%]) developed COVID-19 infection following exposure to unrecognised patients with COVID-19.

**Conclusions:** Unrecognised infections among staff may be a significant driver of HCW infections in healthcare settings. Control measures should be implemented to prevent acquisition from other staff as well as patient-staff transmission.

## INTRODUCTION

The coronavirus disease 2019 (COVID-19) pandemic has introduced significant challenges to the safe provision of healthcare. Healthcare workers (HCWs) are essential for the care of patients infected with COVID-19 and thereby place themselves at risk of acquiring COVID-19.^1^ In a recent report of HCW COVID-19 infections in Victoria, Australia, the state Department of Health and Human Services (DHHS) reported that 72.9% of COVID-19 infections in HCWs were acquired in a healthcare setting.^2^ High rates of HCW infections have been attributed to several factors, including inadequate personal protective equipment (PPE), exposure to large numbers or a high density of COVID-19 infected patients, poor ventilation, worker fatigue and limited access to diagnostic tests.^3-8^ At our hospital, we observed an increase in HCW COVID-19 infections over a two-week period in July and August 2020 which predominantly involved staff working on a single ward. Here, we describe the staff infections at our institution and the interventions introduced to prevent subsequent transmission.

## METHODS

### Setting

Austin Health is a tertiary hospital in Victoria, Australia that operates >900 beds and includes state-wide transplantation and clinical specialty services. During the pandemic, several wards were repurposed as dedicated areas to cohort patients with COVID-19 and were designated “COVID wards”. Maximum patient occupancy for COVID wards was reduced from standard capacity to minimise density quotients for shared rooms, while maintaining sufficient staffing to accommodate the increased time requirements for changes of PPE and additional cleaning. Use of PPE was in accordance with state (DHHS) guidelines^9, 10^ and included a “PPE spotter” on COVID wards. Routine cleaning of COVID wards was performed in line with national guidance^11^ and included daily cleaning and disinfection using Chloradet™ (Agar Cleaning Systems, Preston, Australia) with additional cleaning of frequently touched surfaces. All staff, visitors and patients were screened for symptoms of COVID-19 upon entry. Staff who returned a positive test to SARS-CoV-2 were required to immediately notify the Infection Prevention and Control team, the Occupational Health and Safety team, and their local manager. Samples were taken according to state and national guidelines^12, 13^ using a combined throat and deep nasal swab. Positive test samples or an aliquot of extracted nucleic acid from positive samples were sent to the public health laboratory for genomic testing and analysis using methods previously reported.^14^

### Data Collection

All Austin Health staff who were notified to the organisation after returning a positive test by polymerase chain reaction (PCR) testing for SARS-CoV-2, irrespective of symptoms, were included in the analysis. Data from contact tracing interviews conducted at the time were included in the study with additional details sought retrospectively.

### Definitions

HCWs were defined as staff who had a clinical role in directly caring for patients i.e. those who had physical contact or were in close proximity (<1.5 metres) to patients. Cases were attributed to a particular source based on the criteria outlined in Supplementary Table 1. Contact definitions were taken from DHHS guidelines available at the time^12^ and adapted for local implementation to account for the routine use of PPE by staff (Supplementary Figure 1). All staff who had unprotected exposure to confirmed cases of COVID-19 were required to be tested for SARS-CoV-2 at baseline and day 11 after last exposure to an infected individual, with additional testing performed if they became symptomatic.

### Statistics

Statistical analysis was conducted in RStudio (R version 4.0.3). Confidence intervals for sample proportions were calculated by exact binomial distribution. Incidence rates of HCW infection for a specified group (e.g. ward or HCW type) were standardised to cases per 100 patient exposure days, where the number of patient exposure days was calculated by:

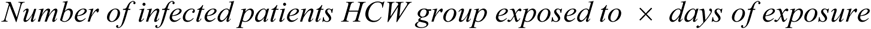

### Ethics

This study was approved by Austin Health Office for Research (CD 20/011 and Audit/20/Austin/169).

## RESULTS

Between 25 January 2020 and 25 November 2020, a total of 49 staff recorded a positive PCR result for SARS-CoV-2 among 8327 staff working at Austin Health (Figure 1). Of these, four staff were excluded from the analyses after an initial indeterminate result and negative results on subsequent testing. There were 45 staff infections included in the analysis, with 28/45 (62% [95% CI 47–76%]) presumed to be healthcare acquired (19 Austin Health; 9 other) on the basis of the source attribution (Figure 1 and Supplementary Table 1). Of the 19 infections likely to have been acquired at Austin Health, 10/19 (53%) involved staff working on a dedicated COVID ward (“Ward A”), clustered in time around early August 2020 (Figure 2).

**Figure 1:**
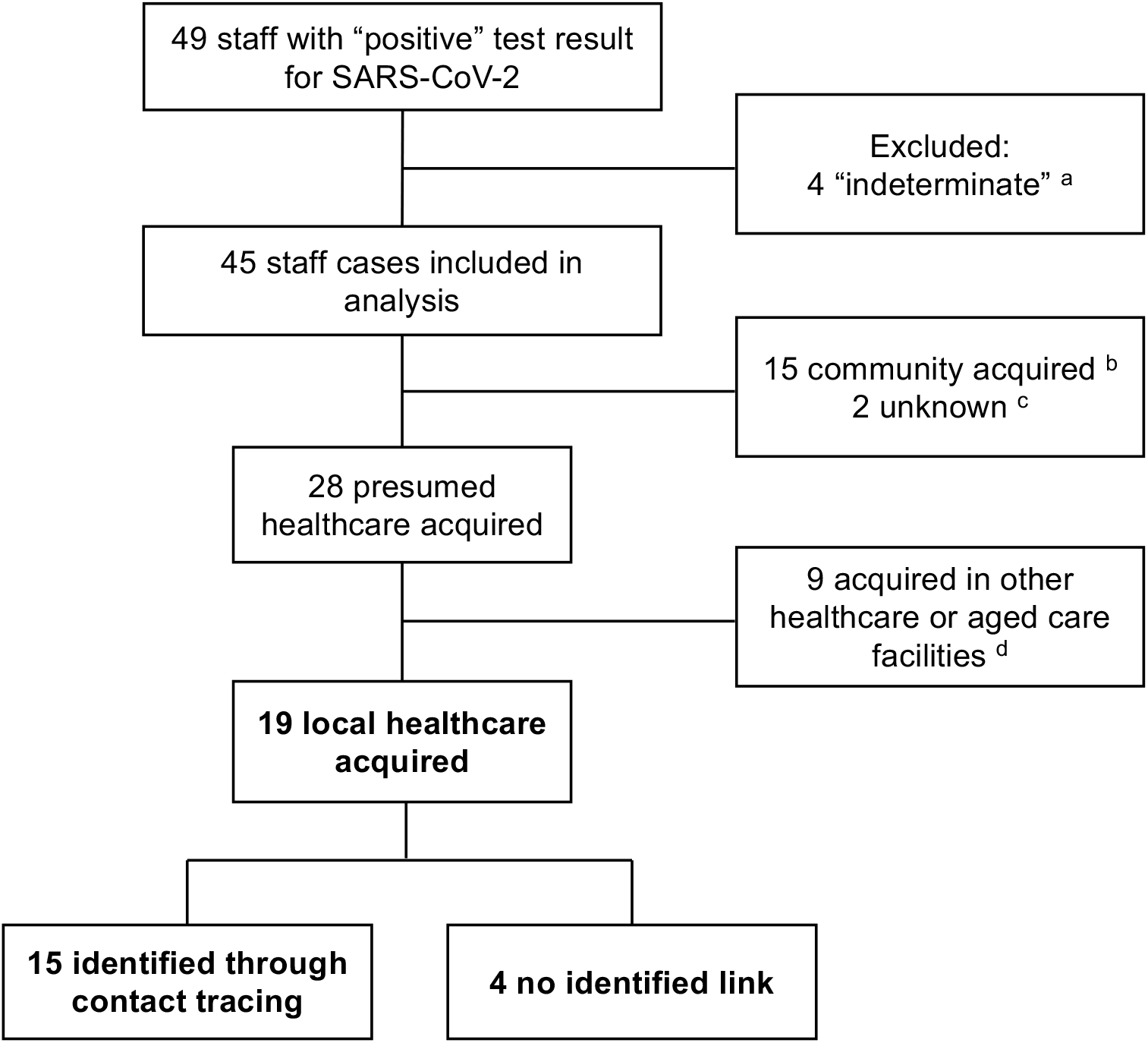
Attributed source of infection for health care workers infected with COVID-19. ^a^ 4 results were initially “indeterminate” but were negative on repeat testing of the same sample and subsequent samples ^b^ Includes 9 cases due to exposure to a household contact, 1 case related to overseas travel, 4 cases unknown exposure but who were working from home and had not been on site ^c^ No healthcare contact with confirmed or suspected cases; unknown community exposure or exposure in other healthcare facilities ^d^ Includes 4 staff who did not work at Austin Health in the 2 weeks prior to their infectious period, 4 staff who cared for patients with COVID-19 at other healthcare facilities but not at Austin Health, and 1 staff member who cared for COVID-19 patients in precautions at Austin Health but was tested as part of an outbreak investigation involving multiple staff and patients at another healthcare facility.

**Figure 2:**
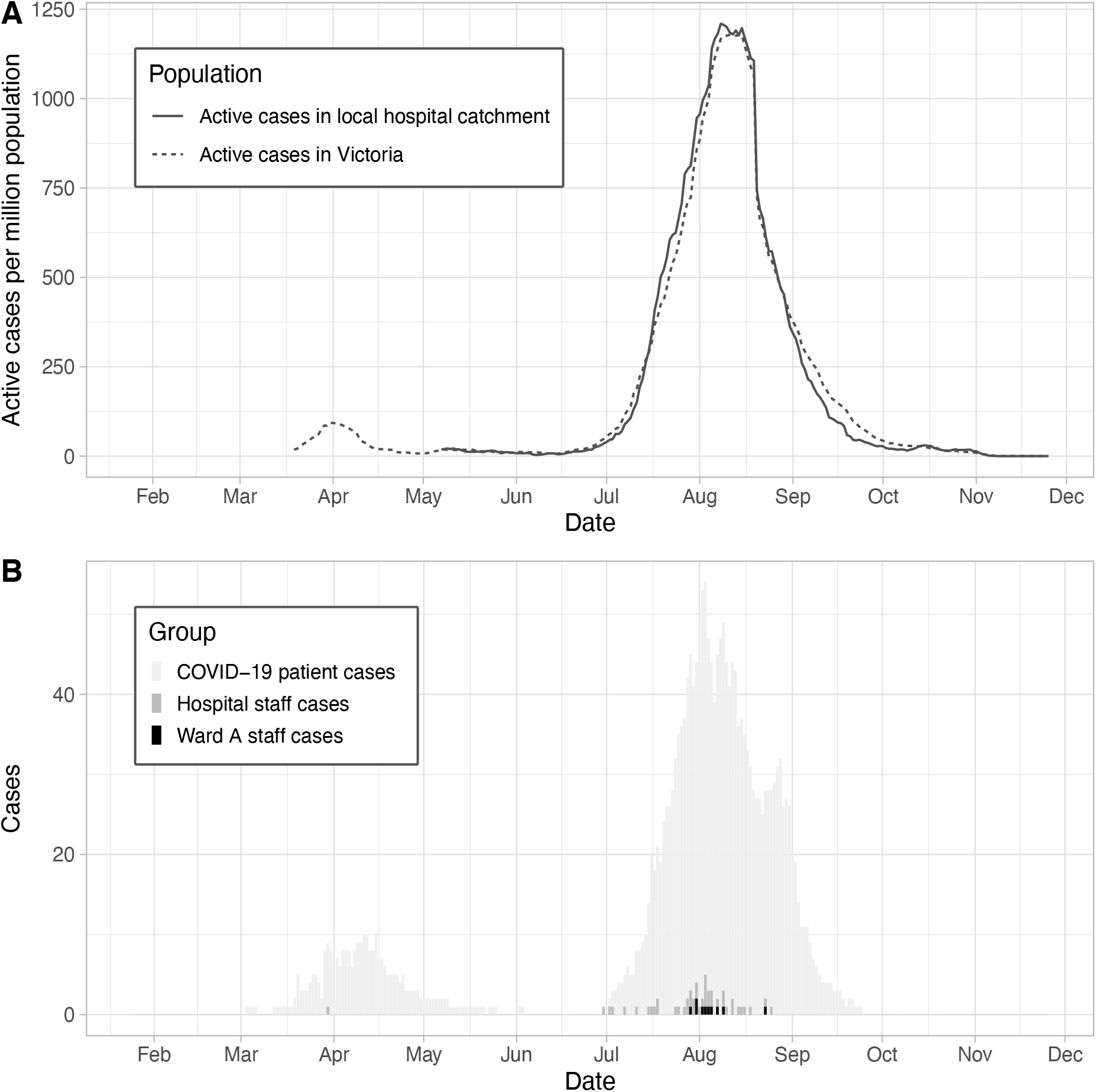
**A**. Community prevalence of active COVID-19 cases. **B**. Incidence of Ward A and Austin Health staff cases in the context of the daily number of inpatients with active COVID-19 infection.

### Outbreak among staff working on a dedicated COVID ward

Ward A is a 32-bed general hospital ward repurposed in March 2020 to provide care to patients with confirmed COVID-19, including eight single occupancy rooms, and capacity reduced from 4 to 1–2 patients per room in six shared patient rooms. The ward’s Heating, Ventilation and Air Conditioning (HVAC) system was modified to supply 100% fresh air intake and exhaust return air from the ward outside the building to avoid recirculation. Air flow was adjusted to achieve a net negative pressure differential of 3-4 Pa to establish inward air movement to the ward from adjoining corridors. At the time of the detection of the first staff case on Ward A, the PPE worn by staff entering Ward A included P2/N95 respirators (a change made prior to the formal updates in state and national guidelines in response to increasing HCW cases at other healthcare institutions and aligned with the decisions at those institutions at the time^1, 9^), face shield, isolation gown, and gloves. Most nursing and cleaning staff worked exclusively on Ward A and were experienced in caring for patients infected with COVID. Ward A shared the same PPE donning room and staff bathroom with another adjacent COVID ward (Ward B), but had separate doffing areas (located immediately prior to exiting each ward) and dedicated staff tearooms outside the clinical areas.

The first case identified on Ward A returned a positive PCR test to SARS-CoV-2 in late July 2020 after developing mild respiratory symptoms (Figure 3 and Table 1). Testing of contacts identified Case 2, who had tested negative on two occasions in the previous week after exposure to a positive HCW on another ward in mid July. Case 2 acknowledged having respiratory symptoms when interviewed with an interpreter, and had worked six shifts during the infectious period. All staff who had worked on Ward A during the infectious period were tested upon detection of Case 2, and again at day 7 and 11 after last exposure on the ward. Additional upstream source contacts were also identified and tested. Due to the likelihood of secondary and possibly tertiary transmissions, all staff that worked the same, preceding or following shifts on Ward A as either Case 1 or Case 2 during the infectious period were furloughed. The case load of Ward A was reduced to 8 patients to accommodate for decreased staffing.

**Table 1:** Characteristics of confirmed cases with COVID-19 infection who worked on Ward A.

| Case | Primary Ward | PPE breach reported | Contact category | Recalled tearoom exposure <sup>a</sup> | Community prevalence <sup>b</sup> |
| --- | --- | --- | --- | --- | --- |
| 1 | A | No | Moderate | Yes | 594 |
| **2** | **A** | **Yes <sup>c</sup>** | **NA** | **NA** | **202** |
| 3 | A | No | Casual | No | 1,747 |
| 4 | A | No | Moderate | Yes | 693 |
| 5 | A | No | Moderate | Yes | 1,098 |
| 6 | A | No | Close <sup>d</sup> | No | 2,240 |
| 7 | A | No | Moderate | Yes | 2,050 |
| 8 | C <sup>e</sup> | No | Moderate | Yes | 2,069 |
| 9 | A | No | Moderate <sup>f</sup> | Yes <sup>f</sup> | 2,527 |
| 10 | A | No | None | No | 751 |

**Figure 3:**
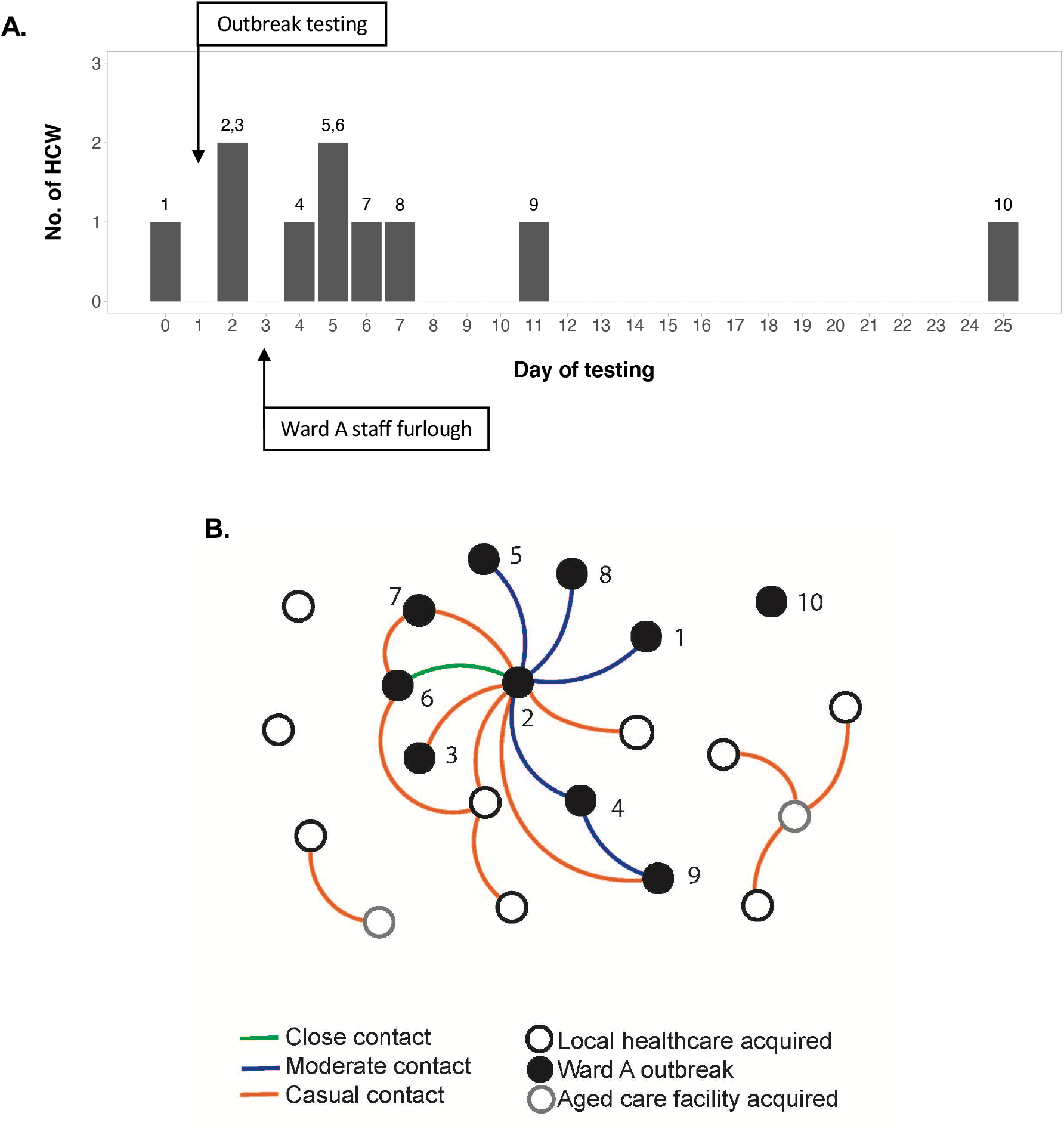
COVID-19 infections acquired at Austin Health. **A**. Timeline of HCW COVID-19 infections in the Ward A outbreak. Numbers above each column indicate the corresponding case number of the HCW (see Table 1). **B**. Staff-staff linkages among staff diagnosed with COVID-19. HCWs involved in the Ward A outbreak are shown in black and numbered, with Case 2 presumed to be the primary case. Linkages are coloured based on contact assignment through contact tracing following each exposure. Case 10 was identified while the Ward A outbreak was still considered “active”, but was not thought to be linked to the other staff. Two additional HCWs (shown in grey) who also worked in aged care facilities with active outbreaks have been included as the initial presumed source for subsequent staff infections.

In total, 179 HCW contacts exposed to Cases 1 and 2 were interviewed as part of the outbreak investigation, identifying 11 close contacts, 82 moderate contacts and 86 casual contacts. 95 HCW contacts had worked on Ward A during the infectious period and provided care to patients with COVID-19. Of these, close and moderate contacts predominantly comprised staff who recalled sharing the Ward A staff tearoom with Case 2. 45/95 (47%) of the Ward A HCW contacts did not use the staff tearoom at the same time as Case 2, and included medical and allied health staff who worked the same hours on Ward A but who used different break facilities, and night-shift nursing staff who did not work the same or overlapping shift hours. None of these staff returned a positive test. Of the other 50 Ward A HCW contacts, seven returned a positive test for SARS-CoV-2 (7/50; 14% [95% CI: 6–27%]; vs 0/45, *p* = 0.01, Fisher’s exact test). The incidence of positive HCWs on Ward A was 1.2 per 100 patient days of exposure compared to 0.7 per 100 patient days of exposure among HCWs on other COVID wards (incidence rate ratio = 1.76; [95% CI 0.64–4.84]). Genomic analysis indicated all cases were derived from the same D.2 lineage (formerly B.1.1.25), as were the majority of cases in Victoria at the time.^15^

Case 8 returned a positive result during the same period, but worked on a different ward and was not initially linked through contact tracing. Although this HCW had cared for patients with suspected COVID-19, the HCW did not have any contact with patients with confirmed COVID-19. However, the HCW reported having meal breaks in the Ward A tearoom during the exposure period with colleagues who worked on Ward A, and was subsequently linked to the Ward A outbreak.

In response to the outbreak, staff break room policies were revised to apply stricter controls to mitigate the potential risk of transmission (Table 2). Following the return to work of HCWs after furlough, ward occupancy was increased again to capacity with patients with COVID-19. One additional HCW (Case 10) returned a positive test 14 days after the last positive case, but did not have any contact with any of the other Ward A staff cases during their respective infectious periods. Routine testing of HCWs working on COVID wards was introduced in early September in line with DHHS recommendations. The outbreak was considered resolved in late September, 28 days (two incubation periods) after the last staff member tested positive.

**Table 2:**
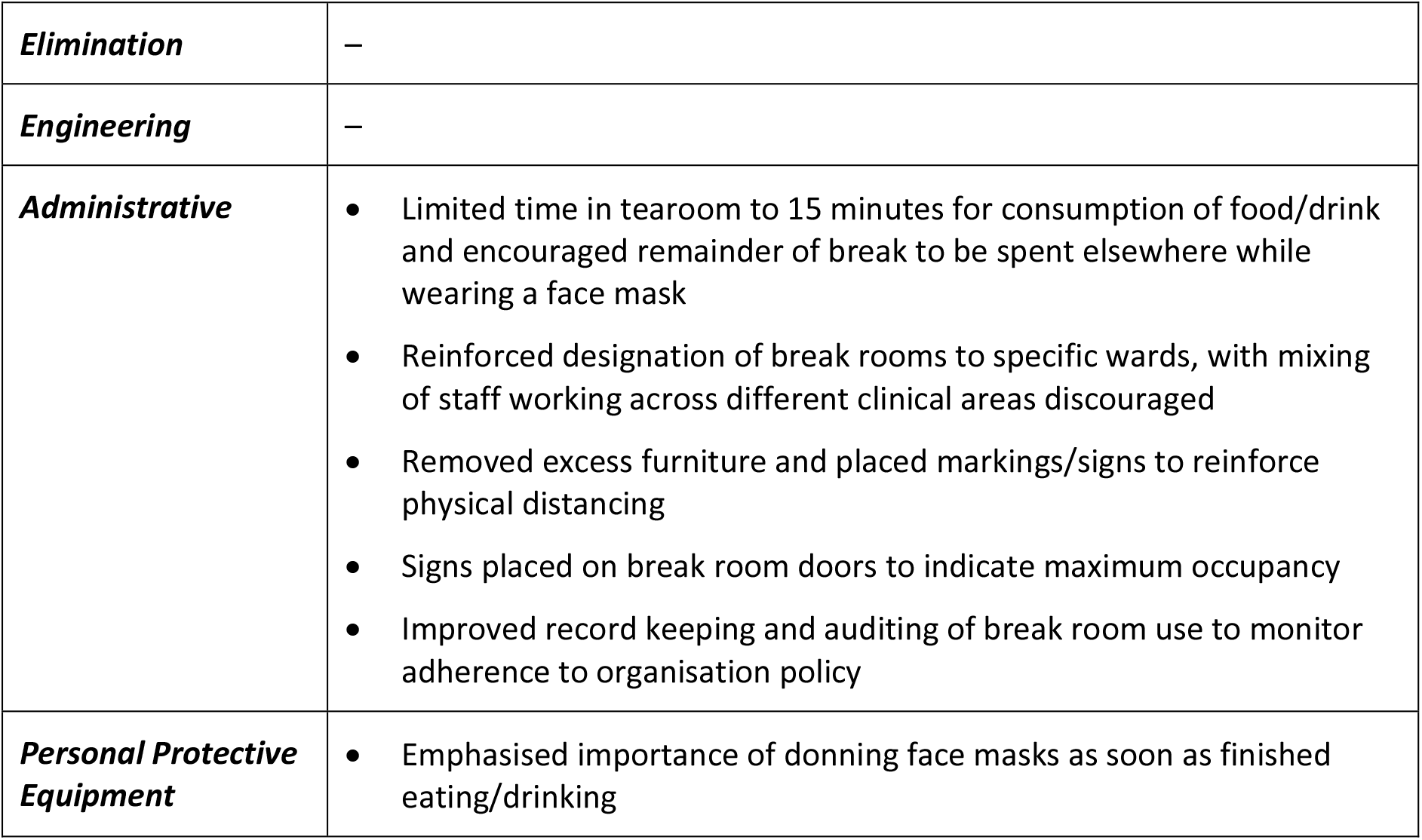
Additional measures implemented to control the spread of COVID-19 in staff break rooms.

|  |  |
| --- | --- |
| ***Elimination*** | – |
| ***Engineering*** | – |
| ***Administrative*** | • Limited time in tearoom to 15 minutes for consumption of food/drink and encouraged remainder of break to be spent elsewhere while wearing a face mask<br>• Reinforced designation of break rooms to specific wards, with mixing of staff working across different clinical areas discouraged<br>• Removed excess furniture and placed markings/signs to reinforce physical distancing<br>• Signs placed on break room doors to indicate maximum occupancy<br>• Improved record keeping and auditing of break room use to monitor adherence to organisation policy |
| ***Personal Protective Equipment*** | • Emphasised importance of donning face masks as soon as finished eating/drinking |

### Other staff cases

Of the other nine staff cases presumed to be acquired at our institution, eight were HCWs. The non-clinical staff member likely acquired COVID-19 when using the same office as a HCW who had returned to work after completing a secondment assisting with the outbreak response at a residential aged care facility. This HCW, who presumably acquired COVID-19 while on secondment, had used the office for non-clinical duties two days prior to developing symptoms, and had vacated the office immediately prior to handing over to the non-clinical staff member.

In addition to the last HCW case on Ward A, three other HCW cases did not have an identified link and were presumed to be independent acquisitions from infected patients (4/19; 21% [95% CI: 6–46%]), including one who reported a clear PPE breach. The remaining five HCW cases outside of Ward A were identified from testing of contacts after investigation of staff cases, and were presumably linked to these other staff.

### Staff exposures to undiagnosed patients

From 25 January 2020 (first case identified in Victoria) until 24 November 2020 (last active case in Victoria deemed non-infectious), 204 HCWs were identified as contacts of patients who were later diagnosed with COVID-19 due to exposure during the infectious period, prior to diagnosis and implementation of appropriate precautions (median exposure = 1 day; range 1–6 days; total 223 patient exposure days). Of these 70 close, 25 moderate, and 109 casual contacts, none (0/204; 0% [95% CI 0–2%]) developed COVID-19 infection from their exposure.

## DISCUSSION

HCW have been disproportionately affected in the number of detected cases in Australia, particularly in Victoria,^2, 16^ though the exact mechanisms for acquisition of infection remain difficult to prove. While the majority of HCW infections are presumed to be acquired at work while undertaking clinical activities, several aspects of our data suggest that these are not necessarily all derived from exposure to infected patients, but may also be driven by exposure to other infected staff working in the healthcare setting. Firstly, the incidence of infected HCWs was clustered both in time and in geographic location. Of the 19 HCWs who possibly or probably acquired infection at our institution, half of these were linked to a single ward. Although this ward cared for patients with confirmed COVID-19 infection, the rate of HCW infections was greater than on other wards that also cared for patients with COVID-19, when standardised for exposure to patients with COVID-19. Nine of the 10 HCW infections linked to the ward occurred within a 2-week period (i.e. a 5% time window of the 9 months that HCWs working on this ward had been caring for patients with COVID-19). Secondly, the involvement of Case 8 in the cluster, who had no contact with patients infected with COVID-19, but who used the Ward A staff tearoom, points towards transmission in this space. In contrast, staff working on Ward A who did not use the tearoom did not test positive in the outbreak investigation. Thirdly, the incidence of HCW infections significantly reduced after changes were made to the protocols around use of staff tearooms, without any change in PPE, ventilation or patient occupancy of the COVID ward.

In Australia, there has been considerable attention focussed on measures to prevent transmission of SARS-CoV-2 from infected patients to staff.^17, 18^ However, as others have also identified internationally, transmission between staff in non-clinical areas can also be a significant driver of HCW infections.^19, 20^ Measures to prevent transmission between staff are critical, given that infected individuals may be infectious despite being asymptomatic or pre-symptomatic,^21^ and staff are generally less suspicious of other staff working regularly in close proximity to them than patients or members of the public. The presence of undetected COVID-19 infection in HCWs was highlighted in a large seroprevalence survey of HCWs who routinely cared for COVID-19 patients, where 6% had serological evidence of previous infection, of whom 29% were asymptomatic in the preceding months and 69% had not previously received a diagnosis.^22^

There are limitations to our retrospective observational data, with only a small number of HCW infections at a single healthcare institution to draw inferences from that may not be representative of the transmission occurring in other facilities. However, our experience provides important local data in recognising the potential for transmission between staff, as has occurred in other essential non-healthcare industries such as food production and distribution. Staff tearooms or break rooms pose a particular challenge. In an environment where transmission is unable to be mitigated by PPE, staff are vulnerable to acquisition from other infected staff. In a study of 703 infected HCWs at a single institution, staying in the same break room as a HCW without a mask for >15 minutes and consuming food within 1 m of another HCW were identified as risk factors for infection.^20^ However, tearoom and break facilities are essential to the wellbeing of HCWs, and provide an area to rest and replenish – critical resources to reducing staff fatigue that can also contribute to HCW infections through errors and breaches in otherwise routine practices such as hand hygiene and doffing of PPE.^7, 8^ Space is often limited in HCW tearooms, with physical distancing requirements further restricting use of these facilities.

A greater understanding of HCW infections is essential to minimising the risk to frontline staff caring for patients with COVID-19. As our data indicate, healthcare institutions must consider risks to staff in both clinical and non-clinical settings, and ensure appropriate measures are in place to mitigate these risks.

## Data Availability

n/a

## ACKNOWLEDGEMENTS

We wish to thank and acknowledge the contributions of the staff infected with COVID-19, the clinical ward staff, Infection Prevention & Control team, staff contact tracing and follow up team, and Health Safety & Wellness staff for their roles in assisting our efforts to understand the infections occurring in staff at Austin Health. We also wish to extend our acknowledgements and gratitude to the laboratory staff in Austin Pathology and the Microbiological Diagnostic Unit Public Health Laboratory.

All contributing authors had full access to the data in this study.

## SUPPLEMENTARY APPENDIX

**Supplementary Table 1:** Risk stratification and source attribution for HCWs with COVID-19.

| **<u>Risk stratification for healthcare acquisition</u>** |  |
| --- | --- |
| Probable | Close contact of a confirmed COVID-19 case while working in healthcare (with preceding or synchronous infection)<br>In quarantine or furloughed due to exposure from active outbreak<br>Reported breach in PPE while caring for a patient with COVID-19 |
| Possible | Worked on COVID ward or cared for patients with confirmed COVID-19<br>Worked on a ward with an active outbreak but not quarantined/furloughed |
| Unknown | No known/minimal contact with confirmed COVID cases in the acquisition period |
| **<u>Risk stratification for community acquisition</u>** |  |
| Probable | Close contact of a confirmed COVID-19 case in the community (with preceding or synchronous infection)<br>In quarantine due to exposure from an active outbreak in a community setting |
| Possible | From a community local government area with high prevalence (defined as >200 active cases per million population = DHHS definition when initiating “hot spot” asymptomatic community testing in July 2020)<br>Works in or visited a location with an active outbreak, but not in quarantine<br>A household member in quarantine due to close contact or exposure in an active outbreak |
| Unknown | No known contacts or risk factors<br>From a community local government area without high prevalence (defined above) |

| **<u>Source attribution from risk stratification</u>** |  |  |
| --- | --- | --- |
| **<u>Healthcare acquisition risk</u>** | **<u>Community acquisition risk</u>** | **<u>Presumed source</u>** |
| **Probable** | **Probable** | **Unknown** |
| **Probable** | Possible | Healthcare |
| **Probable** | Unknown | Healthcare |
| Possible | **Probable** | Community |
| **Possible** | **Possible** | **Healthcare** |
| **Possible** | Unknown | Healthcare |
| Unknown | **Probable** | Community |
| Unknown | **Possible** | Community |
| **Unknown** | **Unknown** | **Unknown** |

*Presumed source was determined after considering the Healthcare acquisition risk and the Community acquisition risk*

**Supplementary Figure 1:**
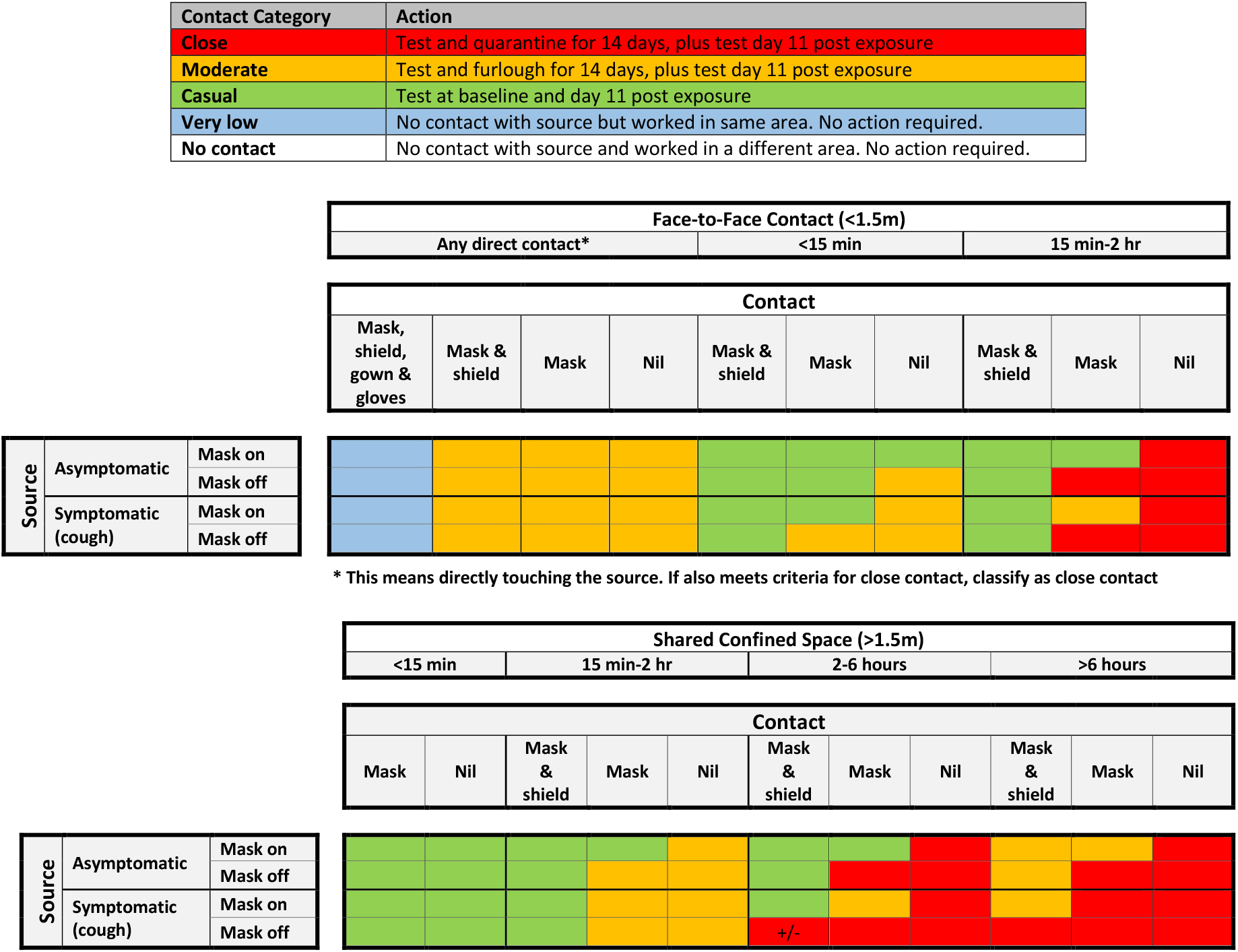
Risk matrix for staff and patient exposures to COVID-19.

